# Cigarette and E-Cigarette Tax Impacts on America’s Oldest Generation of Smokers

**DOI:** 10.64898/2026.01.27.26344945

**Authors:** Jason Semprini

## Abstract

**Background:** As cigarette smoking continues declining among youth and young adults, smoking rates among older Americans remain unchanged. Historically, cigarette and, more recently, e-cigarette tax policies influenced smoking behavior in younger smokers. Understanding how older smokers respond to tax changes can inform public health strategies.

**Methods:** We assembled a quarterly panel of state cigarette and e-cigarette tax rates using the CDC STATE System Tobacco (2000–2024) and E-Cigarette Legislation databases (2015–2024), then merged these data to individual-level survey responses from the Behavioral Risk Factor Surveillance System (BRFSS). Our sample included all adults aged ≥65, but our main specification included adults with a history of smoking. We estimated two-way fixed-effects population-weighted linear probability regression models of current smoking and past-year quit attempts.

**Results:** Among 3,117,382 adults, 50% had a history of smoking; from which 18% currently smoked. A one-dollar tax increase was associated with current smoking: cigarette tax = -0.61-percentage points (CI = -0.94,-0.28); e-cigarette tax = +0.19-percentage-points (CI = 0.14,0.24). There was no association between cigarette taxes and quit attempts. A one-dollar increase in e-cigarette tax was associated with reduced quit attempts (-0.17-percentage-points; CI = -28,-0.06). Approaching tax parity by one dollar was associated with increased current smoking by 0.23-percentage-points (CI = 0.16,0.29) and reduced quit attempts by -0.17-percentage-points (CI = -0.29,0.05).

**Conclusions:** Older American smokers appear responsive to cigarette and e-cig tax changes. Policies increasing the relative cost of e-cigarettes may impede cessation and perpetuate smoking rates in older generations at the highest risk of smoking related harm.

**Implications:** Many studies have investigated the impact of cigarette and e-cigarette tax changes on smoking behavior in youth or younger adults. This study adds new evidence quantifying how new cigarette and e-cigarette taxes change smoking behavior among older adults, a population yet to realize reductions in smoking despite higher risk of tobacco related harm. Analyzing large, population-based survey data, we show that older smokers change smoking behavior due to cigarette and e-cigarette taxes. Taxation of e-cig products may produce unintended harm among older smokers. Specifically, approaching tax parity may increase cigarette smoking and reduce quit attempts among older adults with a history of smoking.

## Introduction

Deemed the forgotten generation, older Americans have not realized the persistent declines in cigarette smoking rates experienced by youth and young adults^1^. For decades, cigarette smoking rates in these younger populations have cut by more than half^1–3^. In older Americans, however, for over 20 years smoking rates have not changed^1^. These reductions in younger groups have been a result of effective public policy and changing social practices. Among the policies most effective at curbing youth or young adult smoking have been cigarette taxes^4–6^. However, evidence suggests that older smokers are less price sensitive than younger populations, potentially limiting the positive effects of raising cigarette taxes^7–9^.

More recently, smoking reductions have been attributed to the rising use of alternative products, notably e-cigarettes^10^. Again, however, although evidence is sparse, older adult smokers appear less inclined to switch to a new-age product like e-cigarettes^10^. Therefore, it is not certain whether the rising popularity of e-cigarettes will spillover into older populations.

Meanwhile, tobacco control efforts have dedicated greater attention to preventing e-cigarette use. Despite evidence that e-cigarettes offer lower relative risk than combustible cigarettes and efficacy as a smoking cessation tool, many states continue implementing e-cigarette taxes^11,12^. Although evidence is clear that these e-cigarette taxes lead to increased smoking in youth or younger adults^4,13,14^, how e-cigarette taxes would affect older adult behavior remains unclear. Such evidence remains critical for public health systems, as approaches to actually drive down the persistent cigarette smoking behavior could dramatically improve health outcomes in this population facing the largest burden of preventable, smoking-related death and disease^15^.

## Methods

### Data and Measures

We assembled a state panel of quarterly cigarette and e-cigarette excise tax rates from the CDC State Tobacco Activities Tracking and Evaluation System for Tobacco (2000-2024) and E-Cigarette Legislation (2015–2024) (https://www.cdc.gov/statesystem). The primary tax measures were (1) continuous e-cigarette tax rates and (2) continuous cigarette tax rates. Using these tax rates, we also created a “parity” variable, measuring the difference between e-cigarette taxes and cigarette taxes. As an alternative specification, a time-variant e-cigarette tax variable was created to measure exposure to any e-cigarette tax.

This panel of tax measures were merged by state and year-quarter to individual-level survey data from the Behavioral Risk Factor Surveillance System (BRFSS) (2000-2024) (https://www.cdc.gov/brfss/annual_data/annual_data.htm). BRFSS is a repeated cross-sectional, phone-based survey covering all 50 states and DC. Our BRFSS sample was restricted to adults aged 65+, with known current smoking status. We had two outcomes, measured as binary variables: 1) currently smoking and 2) attempted to quit smoking in the past year. In addition to analyzing the entire sample, we identified respondents with a smoking history who reported smoking at least 100 cigarettes in their lifetime. BRFSS includes a variety of socioeconomic and demographic variables such as sex, race/ethnicity, age, education, employment, household income, and marital status. BRFSS provides analytic sampling weights to adjust for the complex survey design and yield estimates representative of the population.

### Study Design and Statistical Analysis

Our study employed a quasi-experimental design leveraging within-state changes in cigarette and e-cigarette taxes by quarter and cross-state differences in tax levels. For valid identification, we relied on the assumption that, conditional on covariates and state, year, and quarter fixed effects, any correlation between e-cigarette tax changes and unobserved factors driving smoking behavior did not vary within states over time. This approach and identification strategy have been employed previously in the e-cigarette policy evaluation literature^4,9,14^.

The primary analysis was estimated via a population-weighted linear probability regression model, shown in Equation 1:

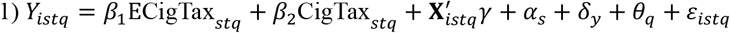

Here, Y_istq_ (binary) denotes either current smoking status or past-year quit attempt for individual = i, in state = s, interviewed in year = t, and quarter = q. ECigTax_stq_ is the state e-cigarette tax rate and CigTax_stq_ is the state cigarette pack rate both measured at the state-quarter level.

Next, we estimated an alternative “tax parity” specification, replacing the two tax variables with a single measure capturing the difference between e-cigarette and cigarette tax rates as shown in Equation 2:

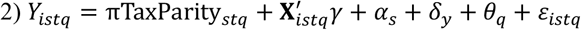

For each equation, X was a vector of dichotomized sociodemographic control variables, α were state fixed effects, δ were year fixed effects, and θ were calendar quarter fixed effects. Robust standard errors were clustered at the state level to account for serial correlation and within-state correlation in the error term induced by shared policy exposure within states. In addition to testing for differential pre-trends with an event-history specification, we conducted sensitivity checks with a binary measure of e-cigarette tax exposure. All analyses were performed in Stata v. 19 using the *reghdfe* package.

## Results

### Summary Statistics

Among 3,117,382 older adults, 48% had a history of smoking and 18% of ever smokers reported smoking currently (Table 1). The sample was mostly female (56.6%), non-Hispanic White (79.1%), held a college degree (53.2%), and married (55.8%). Most respondents were not working (80.1%), with 25.9% of the sample reporting a household income under $25,000. Overall, 8.9% of older adults currently smoked; 17.9% among adults with a history of smoking. Past year quit attempt rates were 3.7% in the full population and 7.8% among ever smokers.

**Table 1:**
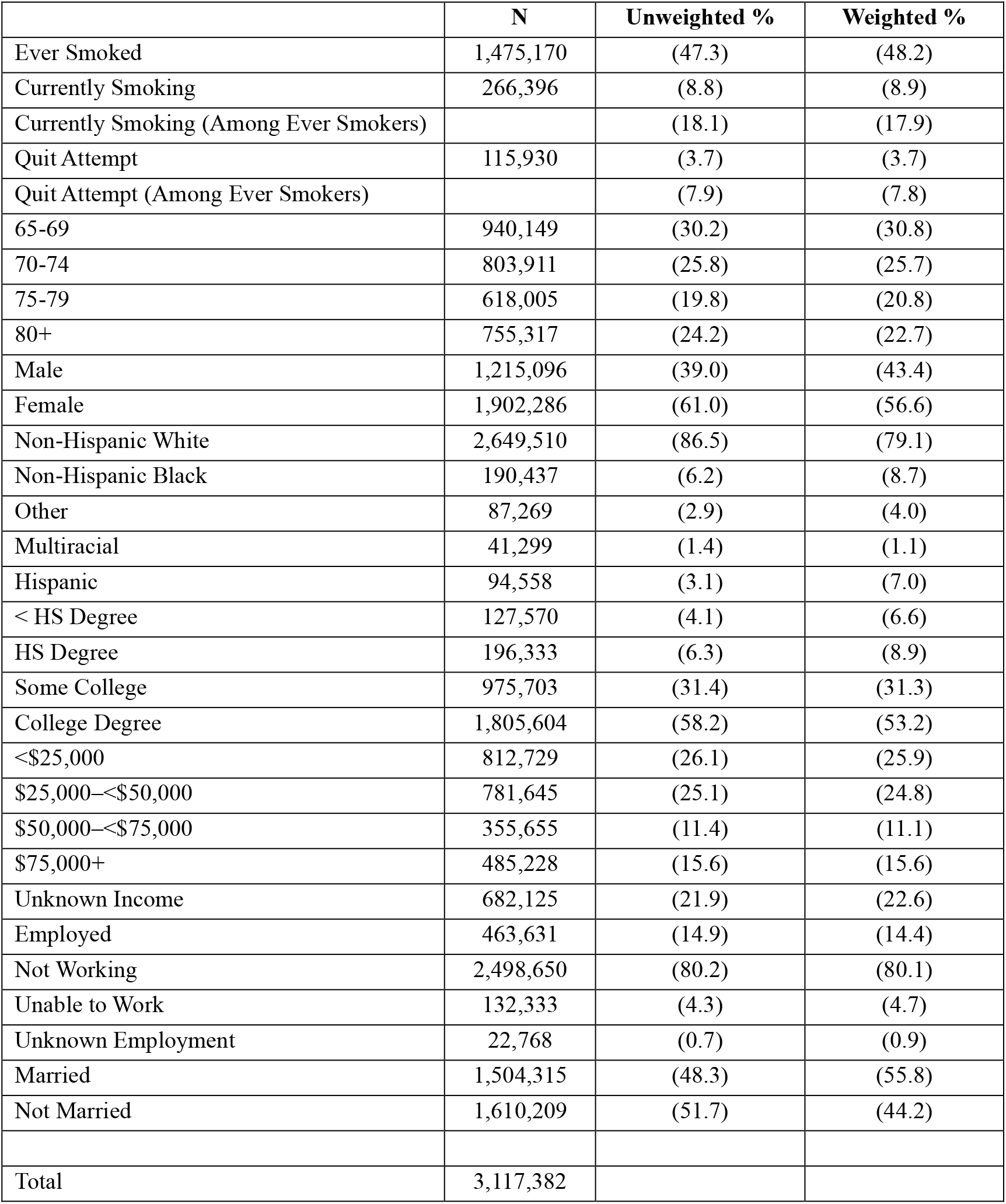
Sample Summary. Table 1 reports sample counts, as well as unweighted and BRFSS survey-weighted percentages for adults aged 65+ in the final analytic sample (N = 3,117,382). “Ever smoked” is defined as having smoked at least 100 cigarettes in one’s lifetime.

**Table 2:**
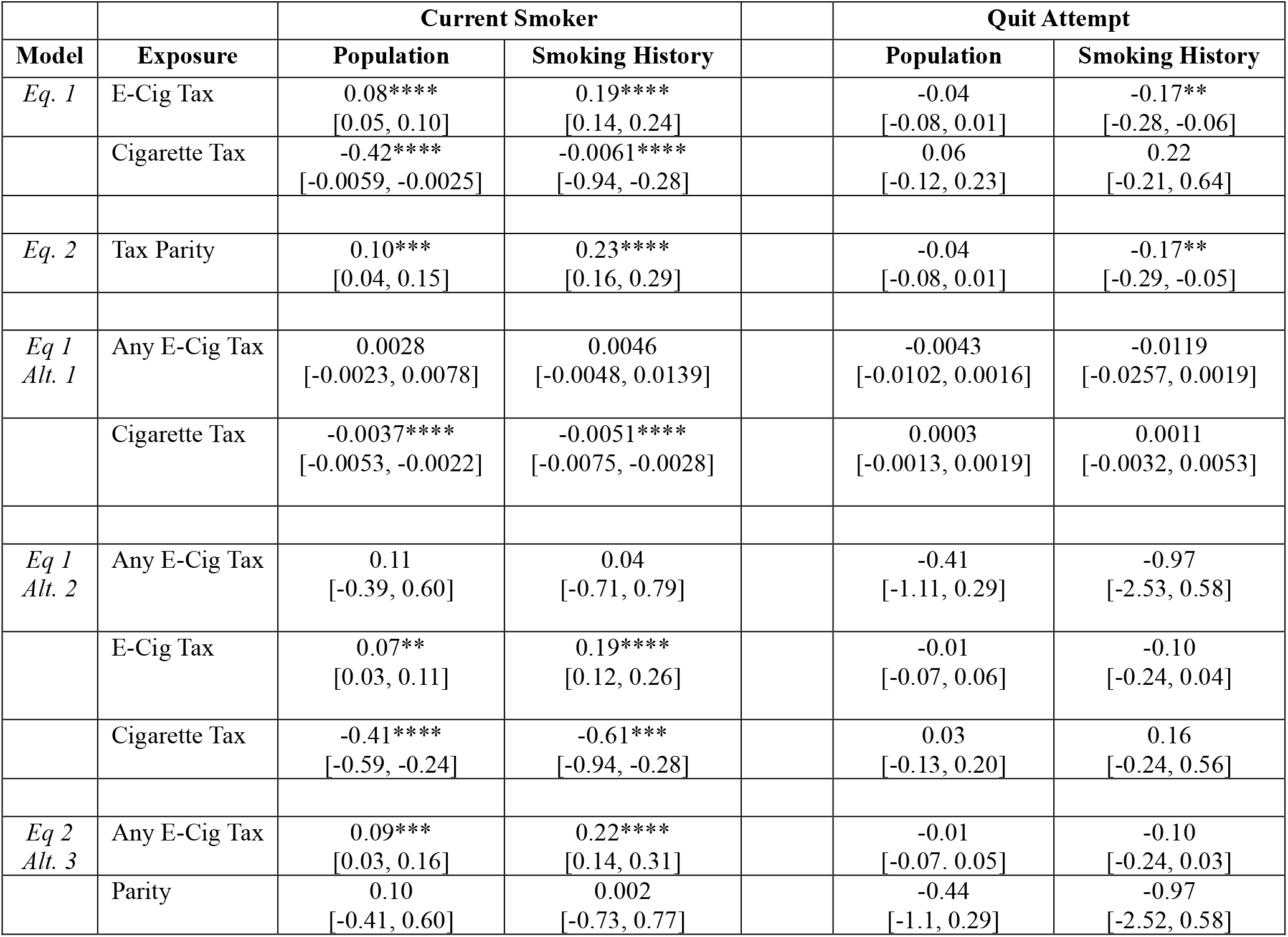
Regression Results. Table 2 reports population-weighted linear probability model estimates (with 95% confidence intervals in brackets) from separate regressions for each outcome and sample. Each model includes the listed exposure variables, adjusts for demographic covariates, and includes state, year, and quarter fixed effects. Robust standard errors are clustered at the state level. When comparing pre-trends in our event history study specification, we found no evidence that smoking rates (Population: p = 0.1413; Ever Smoker: p = 0.1387) or quit attempts (Population: p = 0.1088; Ever Smoker: p = 0.1261) varied between state groups before implementation of e-cigarette taxes. Alt. 1 measures e-cigarette tax as a time-variant binary measure of any e-cigarette tax > 0. Alt 2 includes both a binary and continuous measure of e-cigarette tax. Alt 3 includes both a binary measure of any e-cigarette tax exposure and the e-cigarette-cigarette tax parity continuous measure. Statistical significance is denoted as ** p < 0.01, *** p < 0.001, and **** p < 0.0001.

### Regression Results

Our regression models simultaneously estimated the association between e-cigarette tax and cigarette tax changes on smoking outcomes, among people with a history of smoking (Table 3). We found that a one-dollar cigarette tax increase was associated with a reduction in cigarette smoking (Est = -0.61-percentage points; CI = -0.94, -0.28; p < 0.0001), while a one-dollar e-cigarette tax increase was associated with an increase in cigarette smoking (Est. = +0.19-percentage-points; CI = 0.14, 0.28; p < 0.001). These estimates represent 3% and 1% relative differences from average smoking rates, respectively. Estimates in the overall population were similar (Cigarette Tax: Est = -0.42; CI = -0.59, -0.25; p < 0.0001) (E-cigarette Tax: Est = +0.08; CI = 0.05, 0.10; p < 0.0001).

Next, we found no association between cigarette taxes and quit attempts among the total population (Est. = +0.06-percentage points; CI = -0.12, 0.23; p = 0.502) or among people with smoking history (Est. = 0.22-percentage points; CI = -0.21, 0.64; p = 0.309). Neither did we find any association between e-cigarette taxes and quit attempts in the total population (Est. = -0.04; CI = -0.08; 0.01; p = 0.131). However, among older adults with a history of smoking, we found that a one-dollar e-cigarette tax increase was associated with reduced quit attempts (Est. = -0.17-percentage points; CI = -0.28, -0.06; p = 0.004). This estimate represents a 2% relative difference from average quit rates.

Finally, among older adults with a history of smoking, we found that a one-dollar increase towards e-cigarette-cigarette tax parity was associated with increased current smoking (Est. = 0.23-percentage-points; CI = 0.16, 0.29; p = 0.0004) and reduced quit attempts (Est. = 0.17-percentage-points; CI = -0.29, -0.05; p < 0.0001). These estimates represent 1% and 2% relative differences from average quit rates.

Pre-trend tests found no differences between states by e-cigarette tax status before implementation. See table 1 for sensitivity check results.

## Discussion

While only in existence since 2015 many studies have investigated the impact of e-cigarette tax changes on smoking behavior in youth or younger adults^12^. According to a recent systematic review on e-cigarette taxation, only one study found that older adults responded to e-cigarette tax changes^16^. This study, however, included only two years of data and did not include cigarette smoking behavior as an outcome^16^. The few studies that explicitly investigated how older adults responded to cigarette tax changes revealed modest and mixed effects^8,9^. No studies have modelled how both e-cigarette tax and cigarette tax changes impacted smoking and quit patterns in older Americans. Our study adds new evidence from a decade of e-cigarette tax policy, quantifying how new cigarette and e-cigarette taxes affect contemporary smoking behavior among older adults.

Our results reiterate the potential for cigarette taxes to curb smoking, even in older groups with lower sensitivity to price^9^. Most interestingly, our results suggest that taxing e-cigarettes may produce unintended harm among older smokers. Specifically, approaching tax parity may increase cigarette smoking and reduce quit attempts among older adults with a history of smoking.

Although a larger proportion of 18-24-year-olds smoke cigarettes than those aged 65+, because of US population composition there are three million more smokers in the older age group^17^. Moreover, these older smokers tend to smoke more cigarettes and more frequently, with less desiring or planning to quit^18^. Thus, the declining rate of smokers among younger adults, paired with the rising population of older adults and stagnate smoking rates will result in an even larger distribution of older smokers in the coming decades.

At the individual level, age-tailored smoking cessation programs will prove critical for older adults who smoke cigarettes. But, even if successful, these individual-level targeted programs may not be sufficient to counter the growing population of older adults who smoke. Rather, health systems must proactively implement policies to promote smoking cessation and reduce smoking frequency. Our results reveal that e-cigarette taxation does not appear to do either. In fact, states continuing to seek e-cigarette tax parity with cigarettes may be inadvertently driving older adults to smoke cigarettes.

Extrapolating our estimates to the sixty million Americans over age 65, achieving e-cigarette tax parity would add 120,000 older smokers. Each would likely die a decade sooner than if they had successfully quit smoking^19^. Facing the substantial human and financial costs of adding over one hundred thousand smokers to our Medicare system, policy makers must ignore well-intentioned policies harming this forgotten generation of older Americans and instead lean on effective tax policy as a tool in the fight against smoking.

### Limitations

First, our analyses relied on observational data, which did not measure behavior change within the same individuals overtime. Second, while BRFSS began collecting measures of e-cigarette use, these items were limited, and inconsistently available across states and survey years. This lack of data on our key mechanism limited our ability to examine e-cigarette uptake, substitution, or dual-use patterns among older adults in a way that is comparable over time. Future work should prioritize analyzing data sources with available measures of e-cigarette behavior in older populations. Third, the COVID-19 pandemic plausibly affected smoking and cessation behaviors through multiple channels and may coincide with tax policy changes in ways that are difficult to disentangle^20^. However, by including year and calendar quarter fixed effects, we adjusted for broad temporal shocks, limiting the potential bias from the pandemic. Fourth, our outcomes were self-reported and thus may be subject to misclassification due to recall or social desirability bias. Still, such reporting bias was unlikely to vary systematically with exposure to state e-cigarette tax changes. Fifth, BRFSS did not consistently provide detailed measures of smoking intensity (e.g., cigarettes per day) in recent years across all states or modules, limiting our ability to assess whether tax changes influence smoking reductions. Finally, as with all policy evaluations with non-experimental designs, we cannot fully rule out residual confounding from unobserved, time-varying state factors correlated with tax changes. So, we limited causal claims and all causal interpretations should acknowledge these limitations.

## Conclusion

Older American smokers have not experienced dramatic declines in cigarette smoking rates. While previous research has shown the consequences of cigarette and e-cigarette taxes on smoking patterns in youth or younger adults, less is known about how older adults change smoking behavior once exposed to state tax changes. We found that older adults appear sensitive to price and adjust smoking behavior in the presence of cigarette and e-cigarette taxes. Our primary contribution was identifying how taxing e-cigarette products may produce unintended harm in older adults with a history of smoking. Policies that increase the relative cost of e-cigarettes may impede smoking cessation and perpetuate elevated smoking rates in America’s older generation. These results suggest that reducing the relative cost of e-cigarettes could improve health outcomes among older adults at the highest risk of smoking related harm.

## Acknowledgements

None.

## Data Availability

Statement: Analytic data and code available at corresponding author’s publicly accessible repository [BLIND DURING PEER REVIEW].

